# Treatment Effects of Cholinesterase Inhibitors in Alzheimer’s Disease: a Causal Machine Learning Approach

**DOI:** 10.64898/2026.02.11.26346078

**Authors:** Etienne Dedebant, Mathieu Even, Margaux Törnqvist, Fabien Hauw, Tristan Fauvel, Chloé Geoffroy

## Abstract

**INTRODUCTION:** Treatment response in Alzheimer’s disease (AD) varies substantially across patients, yet no validated frameworks exist to estimate heterogeneous treatment effects (HTE) from observational data while controlling for confounding bias.

**METHODS:** We developed a causal machine learning framework integrating expert-guided causal graphs, complementary HTE estimators, sensitivity analyses, and policy learning. We applied it to cholinesterase inhibitors (ChEIs) in MCI due to AD to patients from the NACC and ADNI cohorts.

**RESULTS:** Analysing 4,049 patients with 12-month and 2,223 with 36-month follow-up, all estimators indicated null or negative long-term ChEI effects on cognitive and functional outcomes, notably on functional measures. ChEIs showed slightly more deleterious effects among men than women.

**DISCUSSION:** This framework provides a methodology for estimating HTE from observational data. It revealed no beneficial responder subgroups, highlighting the challenge of detecting treatment heterogeneity in moderately sized cohorts. This approach can inform treatment selection for other AD therapies including memantine, anti-amyloid agents, and emerging treatments.

## 1 Introduction

### 1.1 The challenge of treatment heterogeneity in Alzheimer’s disease

Alzheimer’s disease (AD) is the most common cause of dementia, affecting over 55 million individuals worldwide [1]. Despite advances in understanding its pathophysiology, including amyloid deposition, tau pathology, neuroinflammation, and synaptic dysfunction, therapeutic options remain limited. Established symptomatic therapies, including cholinesterase inhibitors (ChEIs) and memantine, provide only modest average relief, while recently approved disease-modifying treatments such as lecanemab and donanemab show modest benefits accompanied by significant risks. In all cases, substantial inter-individual variability in treatment response remains poorly understood.

Studies have identified distinct neuropathological subtypes associated with different progression patterns, sex-related differences in neurotransmitter systems, and genetic factors such as APOE genotype influencing disease risk and treatment response [2, 3, 4]. This clinical heterogeneity translates into variable treatment response, with genetic polymorphisms and biological sex involved in differential responses to cholinesterase inhibitors (ChEIs) and monoclonal antibodies [5, 6].

Yet standard efficacy evaluations focus on average treatment effects, obscuring clinically meaningful variation. Recent consensus statements emphasize that failure to account for heterogeneity may explain repeated trial failures and advocate for precision medicine approaches [2, 7]. The field critically lacks robust methodological frameworks to rigorously identify and validate heterogeneous treatment effects (HTE) using real-world data (RWD).

### 1.2 Causal machine learning as a methodological solution

Randomized controlled trials (RCTs), while providing the gold standard for establishing treatment efficacy, face inherent limitations. Restrictive eligibility criteria, limited sample sizes, and fixed follow-up durations constrain their ability to capture the full spectrum of patient heterogeneity. In contrast, RWD captures long-term trajectories across diverse patient populations, offering unique opportunities to investigate treatment heterogeneity at scale. However, leveraging RWD for studying drug effectiveness presents challenges. Treatment assignment in clinical practice is not random but influenced by patient characteristics that also affect outcomes (confounding variables). Incomplete data, measurement error, selection bias, and time-varying confounding further complicate analyses. Traditional predictive machine learning models cannot distinguish correlation from causation and therefore cannot answer the counterfactual question central to treatment decision-making: “What would this patient’s outcome have been under alternative treatment?”.

Causal machine learning (causal-ML) addresses these challenges by combining traditional causal inference methods with flexible machine learning algorithms to estimate individual-level treatment effects, identify responder subgroups, and construct optimal treatment policies from high-dimensional observational data [8]. Recent healthcare applications demonstrate that causal-ML can recover HTE despite complex confounding [8, 9], yet systematic applications to AD treatment remain limited.

### 1.3 Study objectives and rationale for studying cholinesterase inhibitors

This study develops and validates a causal-ML framework to identify which AD patients benefit most from ChEIs using two large US cohorts. Our approach: (1) systematically controls for treatment selection bias using clinical expert input and rigorous statistical methods; (2) evaluates effects across multiple outcomes (cognition, function, behavior) and timeframes (short-term and long-term); (3) provides interpretable decision rules to identify patients most likely to benefit.

We focused on ChEIs as a methodological benchmark for several reasons. First, ChEIs have been extensively studied over two decades, providing a rich foundation for validation. Second, the evidence base exhibits precisely the inconsistency our methods address: RCTs report modest short-term cognitive benefits while observational studies frequently find null or negative long-term associations. Indeed RCTs showed significant cognitive improvements with donepezil over 12-24 weeks in mild-to-moderate AD [10, 11], findings also confirmed by several metanalyses studying ChEIs [12, 13, 14]. However, one observational study, found no significant short-term effect [15].

In contrast, long-term efficacy remains less clear. While some RCTs report sustained benefits for up to two years [16, 17], observational studies, often show minimal or negative long-term effects [18, 19], particularly in MCI populations [20, 21, 22, 23]. This temporal divergence suggests that while ChEIs may offer short-term symptomatic relief, their long-term effects on disease progression are more uncertain. Moreover, meta-analyses note substantial heterogeneity between trials [24, 25], suggesting important unmeasured effect modifiers. Finally, prior literature suggests potential responder subgroups based on sex, APOE genotype, and disease stage, but findings remain inconsistent [6, 5].

Our focus is methodological rather than primarily clinical: to demonstrate that causal-ML can disentangle treatment effects from confounding, detect heterogeneity, and identify responder subgroups rigorously. Once validated with ChEIs where extensive prior knowledge exists, this framework can be extended to treatments with limited evidence or emerging therapies where understanding heterogeneity is critical.

## 2 Methods

### 2.1 Data sources

Our study population included patients from two independent data sources: the National Alzheimer Coordinating Center database (NACC) and Alzheimer’s Disease Neuroimaging Initiative database (ADNI), which both include rich data on AD progression. Before defining our study cohort, we ensured that these two data sources had similar data collection processes and a large overlap in variables useful for our study.

#### National Alzheimer Coordinating Center database

NACC Uniform Data Set (UDS) database sample used for this study includes data of 45,272 patients collected at annual visits at 36 Alzheimer’s Disease Research Centers between September 2006 and August 2016. The data include demographic information, self-reported medication history, medication use, family history of cognitive impairment, clinical characteristics, diagnoses, neuropsychological test battery results, and neuropathological characteristics, using standardized forms and measures. The data also contains APOE genotype, that was available for the majority of the sample (86%). Details of the participant recruitment and data collection procedures in NACC have been described previously [26, 27, 28].

#### Alzheimer’s Disease Neuroimaging Initiative database

ADNI is a longitudinal, multi-center observational study launched in 2004 whose primary objective is to identify and validate biomarkers that track the progression of AD [29, 30] (http://www.adni-info.org). ADNI includes data from 4,536 participants aged 55 to 90, including individuals with normal cognition, MCI, and AD. The dataset includes structural and functional MRI, PET scans (assessing amyloid and tau pathology), cerebrospinal fluid and blood biomarkers, genetic information (such as APOE genotyping), and detailed neuropsychological assessments.

### 2.2 Study Population

All subsequent eligibility criteria and baseline definitions were applied separately to each data source, after which the two resulting study populations were combined for subsequent analyzes.

#### Exclusion Criteria

First, participants with clinical conditions that could confound cognitive trajectories independently of the neurodegenerative process of AD were excluded. Specifically, participants were removed when they presented any of the following diagnoses: Down syndrome, Huntington’s disease, normal-pressure hydrocephalus, central nervous system neoplasm, prion disease, cognitive impairment related to substance abuse, major cardiovascular or systemic comorbidities affecting cognition, essential tremor, significant traumatic brain injury, symptoms consistent with chronic traumatic encephalopathy, vascular dementia, or vascular MCI.

#### Inclusion Criteria

The study population included participants with a diagnosis MCI due to AD, defined by having a global Clinical Dementia Rating (CDR) of 0 or 0.5 [31]. In addition, we restricted the cohort to individuals who had not received any ChEI treatment before their MCI was first diagnosed.

### 2.3 Exposure

Exposure was defined as the use of ChEIs specifically donepezil, galantamine, or rivastigmine. Participants reporting treatment with any of these medications at any follow-up visit after their first MCI diagnosis were classified as ChEI users and assigned to the exposed group. Controls included participants who never reported ChEI use at any visit. Table 3 reports the number of patients in each treatment category, stratified by dataset and by the outcome modeling approach (short-term versus long-term).

### 2.4 Timelines

#### Index date

The index date was defined as the date of visit with an indication of MCI.

#### Baseline definition

For treated patients, we set their baseline as the date of first ChEI treatment. To ensure that the treated and control groups had similar cognitive levels at baseline, we applied the following procedure to compute controls’ baseline. First, we computed in the treated group the median time interval between the first MCI diagnosis and the first ChEI treatment. Then, the control group’s matching baseline date was computed by adding the latter median time interval to the date of MCI visit. To further ensure comparability, we retained only control patients who had a visit within more or less 6 months of this matched baseline date.

#### Censoring

To ensure the treated patients were treated continuously with ChEI, we censored individuals at the first visit where they discontinued ChEI treatment or initiated another cognitive-enhancing medication. In some cases, this resulted in participants having no post-baseline observations available, thereby further reducing the effective sample size.

#### Follow-up

All participants were followed from their baseline visit until loss to follow-up or censoring. Because AD progresses slowly while treatment effects may manifest earlier, we analysed two follow-up horizons: a short-term window to capture early treatment effects, and a long-term window to assess sustained progression. For the short-term analysis, participants were required to have a follow-up visit at 12 months more or less 6 months after baseline; for the long-term analysis, at 36 months more or less 6 months after baseline.

### 2.5 Outcome

#### Outcomes definition

Multiple clinical scores commonly used to characterize AD’s progression and available in both NACC and ADNI were considered as outcomes, namely the Clinical Dementia Rating Sum of Boxes (CDR-SB), Mini-Mental State Examination (MMSE), Montreal Cognitive Assessment (MoCA), Geriatric Depression Scale (GDS), and Functional Activities Questionnaire (FAQ). The CDR-SB ranges from 0 to 18 while all other scores range from 0 to 30. For each patient, these scores were available at multiple study visits, with the baseline visit defined as the reference time point.

#### Long-term Outcome

Long-term effects were summarized by the individual rate of change of each clinical score over the full follow-up period. For each patient and each score, a subject-specific slope was estimated by fitting a linear model to all available observations between baseline and the last recorded visit, using time since baseline as the independent variable. This slope was used as the long-term outcome and reflects the average yearly evolution of the score. The corresponding intercept, representing the estimated baseline score, was retained and later included as a potential confounding variable.

#### Short-term Outcome

Short-term effects were defined to capture early changes while accounting for heterogeneity in follow-up timing. For each patient and each score, the follow-up visit closest to one year after baseline was identified, allowing a tolerance window of more or less six months. The short-term outcome was computed as the change in score between this visit and baseline, normalized by the time elapsed since baseline, yielding a comparable rate of change across individuals.

#### Outcome Harmonization

Because higher values of some scores indicate worse clinical status (CDR-SB, GDS, and FAQ), the corresponding outcomes were multiplied by −1. This transformation ensured a consistent interpretation across all measures, such that higher outcome values systematically reflected clinical improvement.

### 2.6 Variables

#### Preliminary Step

We constructed a unified set of variables based on the union of variables available in NACC and ADNI. Variables present in only one cohort were retained, and missing values in the other cohort were imputed during preprocessing. This strategy maximised the information available while maintaining a consistent feature space across datasets.

#### Selection of Covariates

The variables included in the analysis were classified into three categories commonly used in causal inference to characterize their role in the data-generating process. Confounders were defined as variables influencing both treatment assignment and disease progression and were adjusted to block spurious associations. Instrumental variables were defined as variables that affect the assignment of treatment without directly influencing the outcome, except through treatment, and were considered in the context of alternative estimation strategies. Finally, prognostic variables were defined as variables directly associated with the outcome but not with treatment assignment, and were included to improve the precision of treatment effect estimation.

### 2.7 Design of a Causal Graph

To represent the relationships between treatment assignment, disease progression, and relevant covariates, we designed a causal graph. Establishing this graph is a key step in ensuring the correct identification of causal effects when using causal-ML. To do so we developed a structured questionnaire submitted to a clinician specialized in AD. This questionnaire listed all candidate variables, and for each variable, the clinician was asked to specify whether the variable influences treatment assignment, disease progression, or both.

The responses from the clinician were used to draw a preliminary causal graph, where nodes represented the variables and directed edges represented causal relationships between them. This graph served as the foundation for identifying confounders, which are crucial to avoid confounding bias when estimating treatment effects.

We included additional clinically relevant covariates in the causal graph: the medical center (available in NACC only), to account for heterogeneity across recruitment sites, and baseline clinical scores, which capture disease severity and are strongly related to subsequent disease trajectories, time since the first recorded MCI diagnosis to account for differences in disease duration at baseline. A detailed description of all covariates and their availability across data sources is provided in Table 4.

#### RESEARCH-IN-CONTEXT

1. **Systematic review:** Treatment response in AD varies substantially across patients, yet precision medicine approaches remain limited by the absence of validated frameworks to estimate HTE from observational data. Observational studies face challenges in distinguishing true treatment heterogeneity from confounding by indication, selection bias, and methodological artifacts. No existing frameworks systematically validate whether observed subgroup differences reflect genuine biological heterogeneity or spurious findings.
2. **Interpretation:** Using causal-ML in two large observational cohorts, we found that ChEIs in MCI were associated with null or negative effects on cognitive and functional outcomes at 36-month follow-up. Treatment effects did slightly vary by sex, being more deleterious in men than women. However, HTE analyses did not identify strong effect modifiers among APOE ε4 carrier status, baseline cognitive severity, or other measured patient characteristics. Policy learning identified no beneficial subgroups. These findings suggest that MCI patients with relatively intact cholinergic systems may not benefit from cholinergic augmentation, illustrating that substantial biological heterogeneity does not necessarily produce treatment response heterogeneity. However, these findings can also be due to a too small sample size to detect heterogeneity.
3. **Future directions:** This framework should be applied to AD treatments where identifying responder subgroups could transform clinical practice, particularly memantine and anti-amyloid monoclonal antibodies, where modest average benefits and substantial risks make precision medicine critical. To facilitate widespread adoption, we developed an interactive web-based platform that implements this causal-ML framework, enabling clinical researchers and pharmaceutical industry professionals to explore treatment heterogeneity across diverse populations without requiring statistical expertise. Methodological extensions should incorporate longitudinal biomarker outcomes (amyloid-PET, tau-PET, plasma biomarkers), model time-varying treatment strategies and sequences, and employ target trial emulation to strengthen causal claims. Ultimately, prospective validation through embedded pragmatic trials where predicted individual treatment effects guide treatment assignment will determine whether these methods can improve patient outcomes in real-world practice.

### 2.8 Statistical methods

#### Descriptive Analyses

To assess main clinical difference between the ADNI and NACC cohorts, we computed descriptive statistics. Categorical variables were described as percentages and counts and continuous variables as means along with their standard deviations.

#### Imputation of Missing Values

We addressed missing data in two ways: *(i)* for methods that can naturally handle missing values such as random forests, we kept the data as is; *(ii)* for methods that do not support missing values, such as linear models, we imputed the data before performing the analysis. Imputation was performed using the Multiple Imputation by Chained Equations (MICE) method (using the mice R package) and evaluated by plotting distribution graphs of each variable for imputed and non-imputed groups.

#### Average Treatment Effects and Conditional Effects

*Definition*. The average treatment effect (ATE) summarizes the overall expected difference in outcomes between patients receiving the intervention and those receiving the control. In contrast, the conditional average treatment effect (CATE) estimates this difference taking into account differences in patient characteristics. While the ATE provides a population-level measure of efficacy, the CATE incorporates potential heterogeneity in treatment response. Together, these measures help assess both general and subgroup-specific treatment effects.

As researchers have pointed out towards sex-specific cholinergic difference, we computed ATEs within the subgroups of men and women [6].

##### Identification & Causal Assumptions

The ATE and CATE are formally defined using counterfactual quantities, and are identifiable, i.e., can be estimated from observations, as long as standard causal assumptions are satisfied: Stable Unit Treatment Value assumption (SUTVA), positivity and unconfoundedness, which respectively imply that there are no interferences between units, each unit has a non-null probability of having both treatment assignments, and there are no unobserved confounders [32].

To evaluate positivity, we analysed probability bounds obtained by propensity models, computed standard mean differences (SMD) after and before propensity score weighting, and made joint variable importance plots. In case some patients had too extreme propensity scores, trimming was applied, e.g, patients were removed from the study population.

To evaluate unconfoundedness, we applied a refutation test [33]. This approach introduces a placebo variable generated through random coin flips to simulate spurious associations. Examining the resulting ATE can help ensure that the observed treatment effects are not artifacts of the estimation procedure or data structure.

##### Causal Inference Methods

To estimate nuisance parameters, we used classical parametric models (linear regression, logistic regression) vs. flexible data-driven models (random forests). To estimate ATE, we used the following estimators: G-Formula with linear regression for outcome nuisance estimations, Inverse Probability Weighting (IPW) with logistic regression for propensity score estimation and Augmented Inverse Probability of Weighting (AIPW) with both of the previous nuisance parameters combined, causal forests and the R-Learner using the rlasso estimate. Reference for these estimators can respectively be found in [34, 35, 36, 37]. For CATE estimations, we used causal forests and the R-learner. For estimating ATE within a subgroup, we used causal forests which naturally propose an estimator of the ATE within subgroups using CATEs and an AIPW estimator.

##### Confidence Intervals

95% confidence intervals (CIs) were computed by taking into account variability that arises from the imputation, and the causal analysis itself using a bootstrap procedure [38]. Specifically, datasets were obtained by sampling with replacement the original data, after which imputation was performed on the resampled datasets. Causal analysis was then applied to each of the imputed datasets to estimate the treatment effect, reported as the median of bootstraped ATEs and CIs. For causal forests, we reported the ATE and asymptotic confidence intervals provided by grf R library as well as a the previously described bootstrap approach.

#### Subgroup Identification

A key objective of this study was to identify subgroups of patients for whom treatment with ChEIs may be more appropriate, with the aim of supporting interpretable and clinically actionable treatment decisions. Rather than focusing solely on ATEs, we sought to characterize heterogeneity in treatment response and to derive transparent treatment assignment rules. To this end, we employed a policy-learning framework based on optimal policy trees [39]. After estimating individual treatment effects(ITE) on the training split using causal forests, an optimal treatment policy was learned by maximizing the expected policy value over a restricted class of depth-constrained tree-based decision rules, as implemented in the `policytree` package. ATEs were then computed on a test split on our dataset using the learned policy as treatment attribution. More specifically, the policy value was defined as the population average of the ITEs, multiplied by the sign of the policy assignment (positive for units treated under the learned policy and negative otherwise). The ATE over the population with no policy applied was estimated using causal forests.

## 3 Results

### 3.1 Study population

The resulting study population included 4,049 patients (864 treated by ChEI vs. 3,185 controls) for the short-term follow-up analysis and 2,223 patients (430 treated by ChEI vs. 1,793 controls) for the long-term follow-up analysis (details by database can be found in 3 in Appendix). Table 1 summarizes the impact of each inclusion and exclusion step for both NACC and ADNI, from the initial datasets to the final analytic cohorts.

**Table 1:** Attrition of the study population for NACC and ADNI cohorts.

| Step | NACC | ADNI | Total |
| --- | --- | --- | --- |
| Initial datasets | 45,272 | 4,536 |  |
| Exclusion criteria applied | 43,172 | 4,511 |  |
| Existence of index date (first MCI diagnosis) | 12,616 | 1,616 |  |
| Existence of baseline | 5,926 | 995 |  |
| *Short-term follow-up (12 ± 6 months)* |  |  |  |
| Eligible for follow-up | 3,512 | 803 |  |
| After censoring | 3,273 | 776 | **4,049** |
| *Long-term follow-up (36 ± 6 months)* |  |  |  |
| Eligible for follow-up | 1,846 | 490 |  |
| After censoring | 1,754 | 469 | **2,223** |

### 3.2 Expert-Driven Causal Graph

Figure 6 in Appendix illustrates the causal diagram constructed from NACC variables, incorporating all questionnaire-derived variables together with the additional variables. Notably confounders included variables such as age, cognitive and functional scores at baseline, effect modifiers variables such as gender, educational status, diagnosis of bipolar disorder, PTSD, Parkinson’s Disease and instrumental variables medical history of stroke, epilepsy, hypertension.

### 3.3 Statistical Analyses

#### Descriptive Analysis

Table 2 summarizes baseline characteristics of the cohort used for long-term modelling. Participants from NACC and ADNI were similar in age (on average 75.7 years in NACC and 75.2 years in ADNI) and educational level at baseline (on average 15.7 years of education in NACC and 16.0 in ADNI), while the proportion of male participants was higher in ADNI (on average 47.5% in NACC vs. 61.2% in ADNI). In both datasets, the study population was predominantly White (80.6% in NACC and 93.4% in ADNI). Baseline CDR-Sum was higher on average in ADNI compared with NACC (1.3 vs. 1.8). Substantial differences were observed in biomarker availability, particularly for amyloid status with 95.4% missing values in NACC vs. 60.3% in ADNI.

**Table 2:**
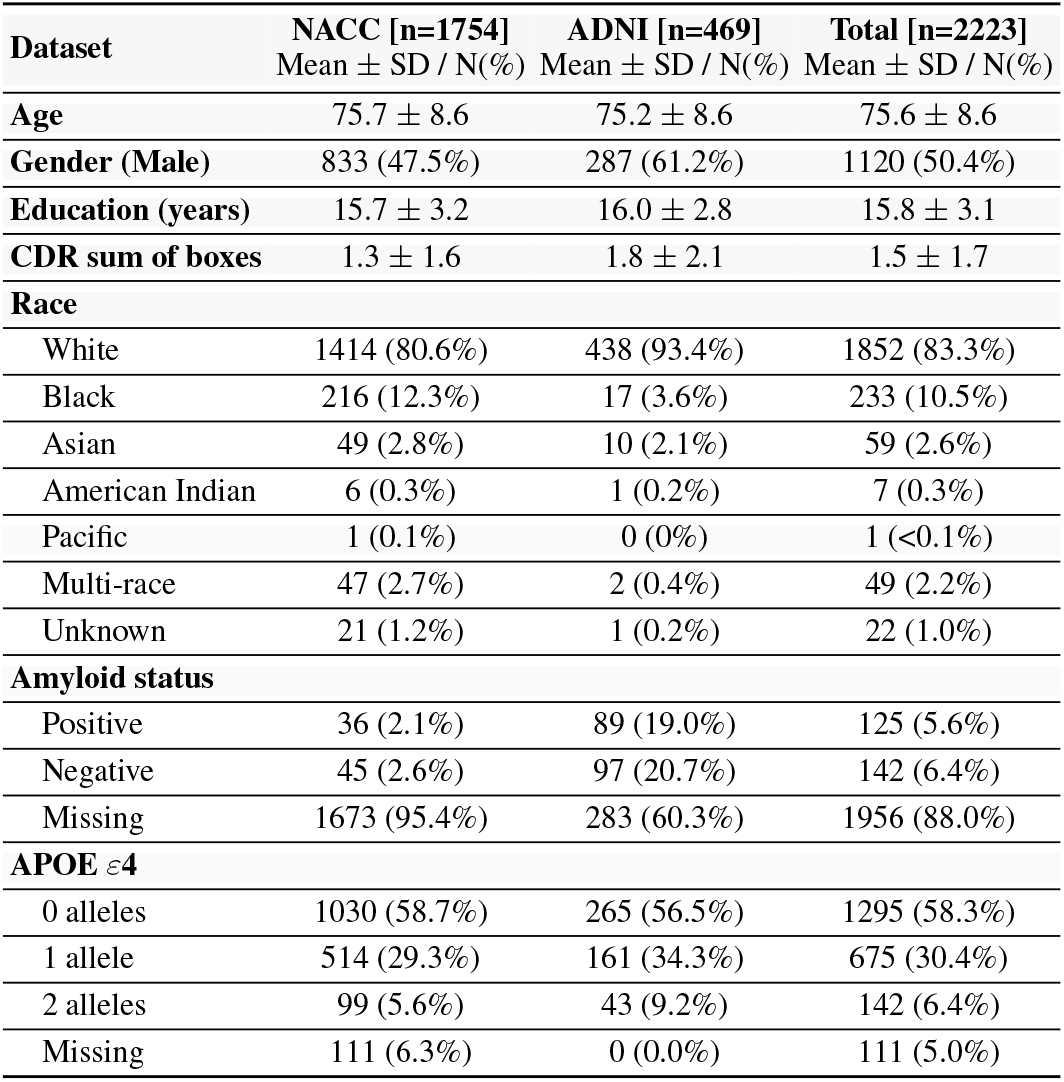
Descriptive statistics of baseline characteristics of the study population used for long-term modeling.

**Table 3:** Treatment group statistics across datasets and modeling horizons.

| Dataset | Short-term modeling |  | Long-term modeling |  |
| --- | --- | --- | --- | --- |
|  | Control | ChEI | Control | ChEI |
| NACC | 2,568 | 705 | 1,379 | 375 |
| ADNI | 617 | 159 | 414 | 55 |
| **Total** | **3,185** | **864** | **1,793** | **430** |

**Table 4:**
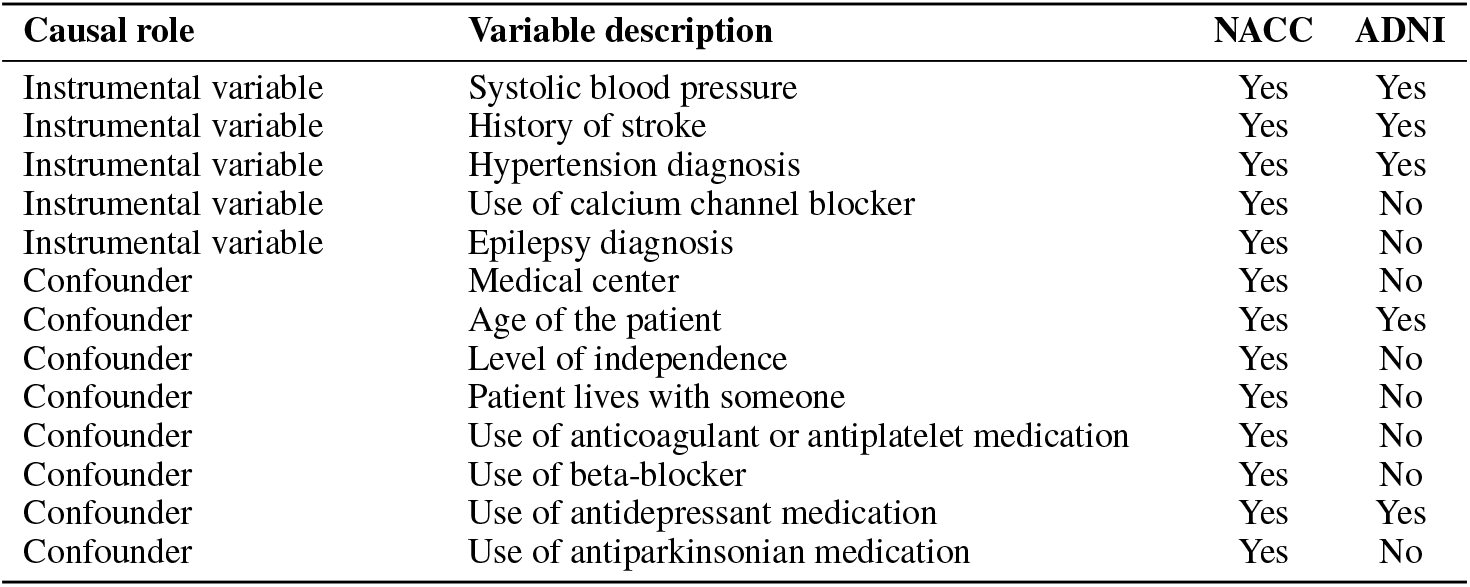
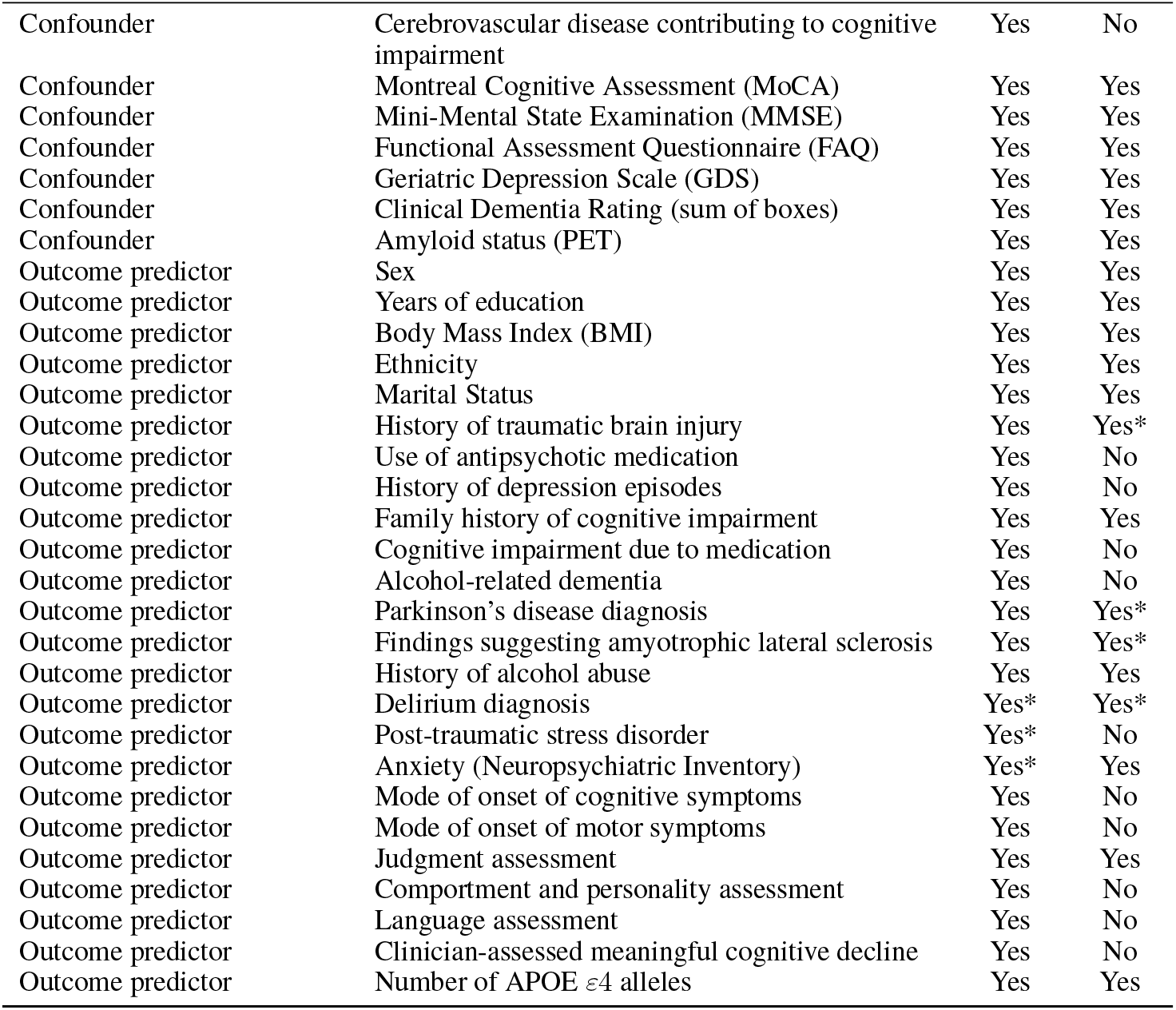
Candidate variables considered for the analysis, grouped by causal role, and their availability across data sources. An asterisk (*) indicates variable that was ultimately excluded due to whether a high proportion of missing values or because it is constant.

#### Imputation of Missing Values

Details on missing values are described in Figure 7 and 8 in Appendix. The high number of variables with 25% of missing values mainly came from variables in NACC missing in ADNI. Overall, comparing distributions of variables before and after imputation showed that our imputation method produced realistic synthetic data (Figure 9 in Appendix is an example on AMYPLET and BPSYS varuables).

#### Causal Assumptions

As described in Section 2.8, we evaluated the positivity assumption by examining the empirical distribution of the estimated propensity scores either obtained with a random forest or logistic regression. Figure **??** shows that for the long-term modeling, a substantial fraction of individuals had propensity scores extremely close to limit values of 0 or 1, indicating a violation of the overlap assumption. Applying trimming with a probability threshold value of *η* = 0.05 resulted in a final population sample of 1944 participants, of whom 429 were treated for the long-term outcome cohort, and 3565 participants, of whom 860 were treated for the long-term outcome cohort).

By design, since we removed patients with a propensity score *η* - close to 0 or 1 for *η* = 0.05, the overlap assumption held (see Figure 1). Figure 1 shows the covariate balancing plot, e.g., love plots, for propensity scores used in ATE computations obtained by fitting a logistic regression or regression forest (for causal forests).

**Figure 1:**
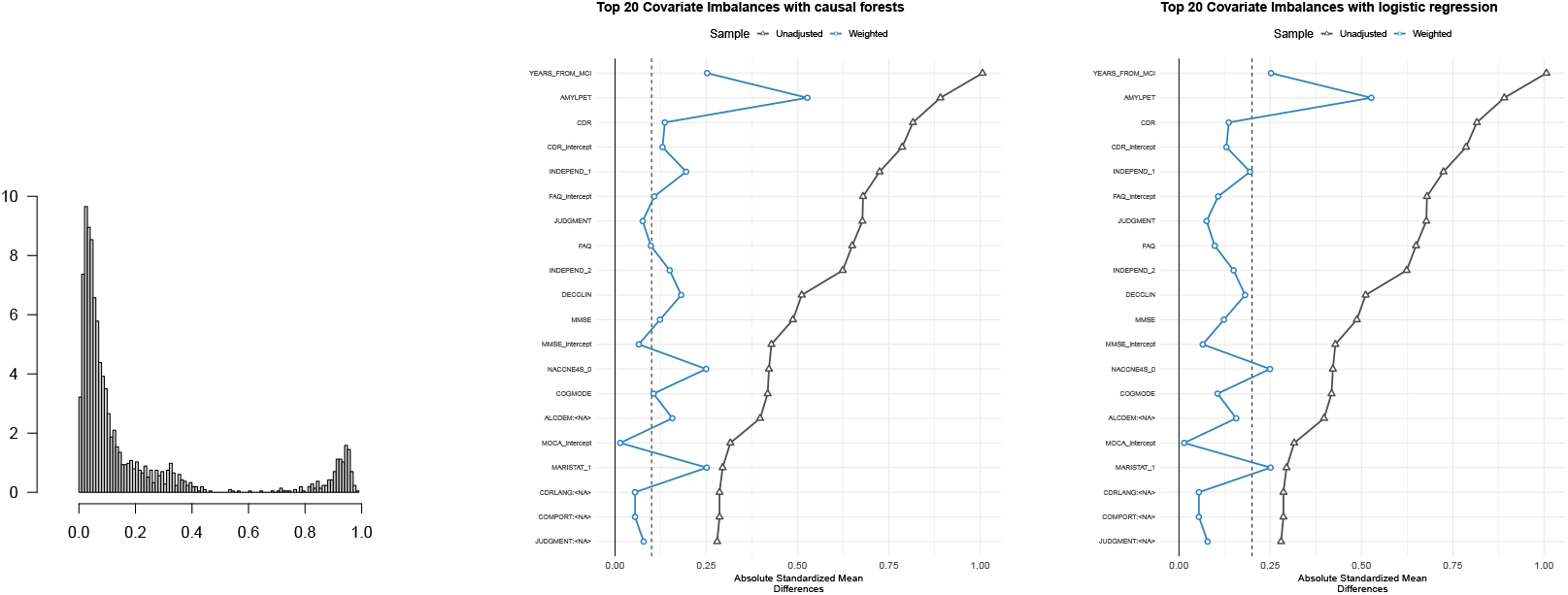
Histogram of propensity scores (left) and love plots for causal forest with a regression forest as propensity model (middle) and for AIPW and R-learner obtained with a logistic regression propensity score model (right).

#### Average Treatments Effects

Figure 2 and 3 show the ATEs computed with the five estimators, on all five different scores, for respectively the long-term and the short-term outcomes.

**Figure 2:**
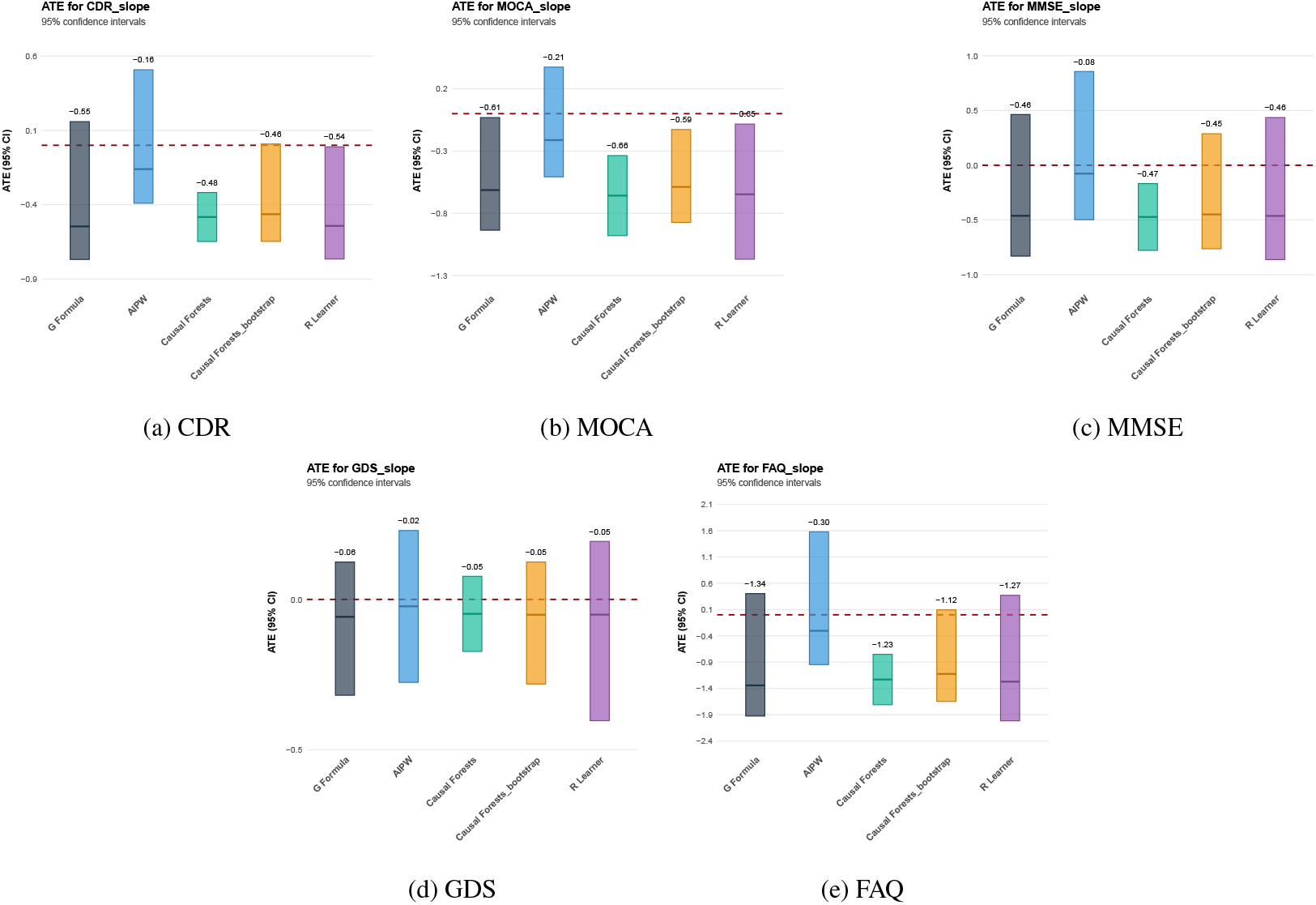
Average Treatment Effects for long-term outcomes estimated across several scores using multiple estimators.

**Figure 3:**
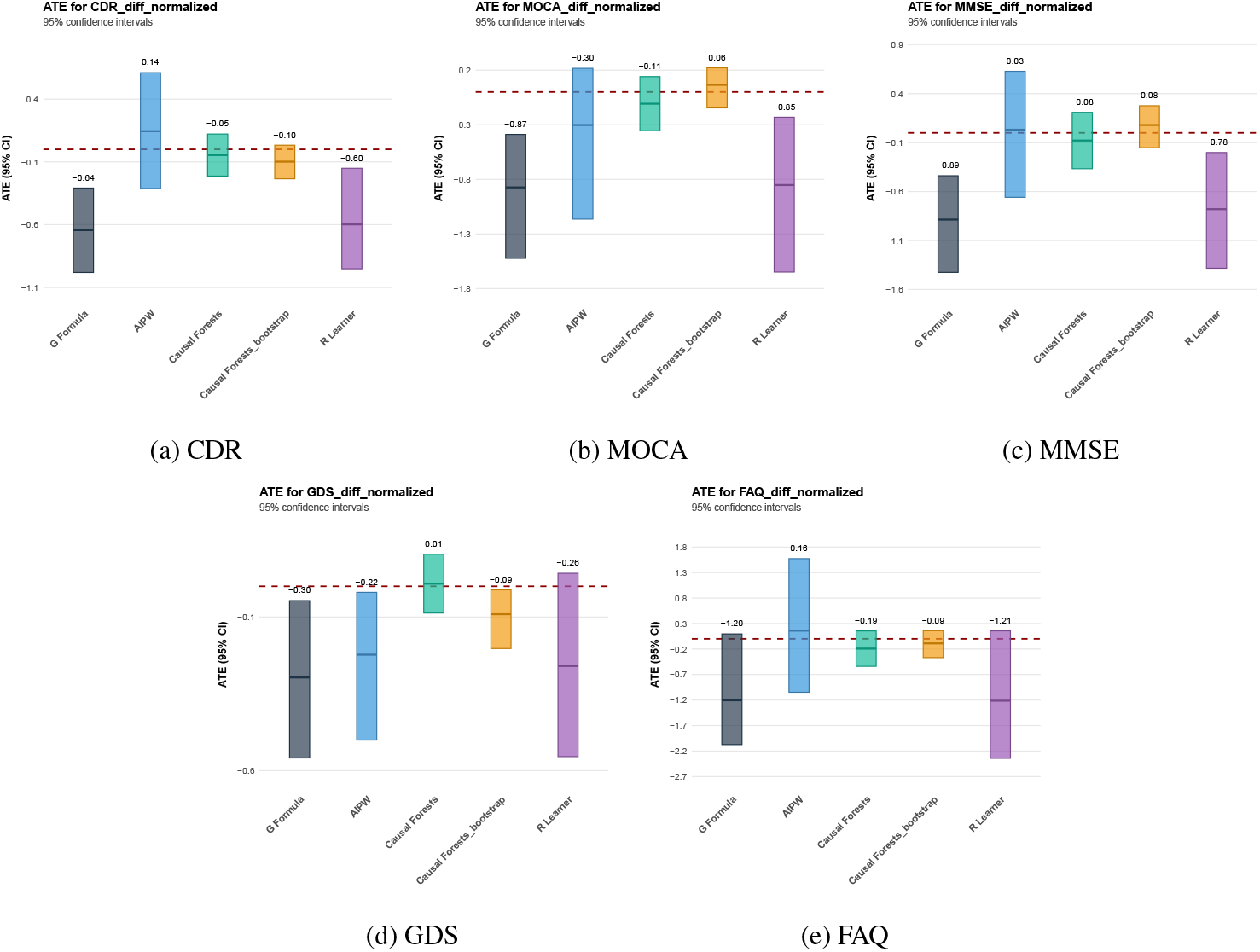
Average Treatment Effects for short-term outcomes estimated across several scores using multiple estimators.

##### Long-term Outcome

For long-term outcomes, we observed a consistently small deleterious effect on cognitive performance, most notably for MOCA, with negative and statistically significant ATEs ranging from −0.61 to −0.69 for 4 out of 5 estimators, whereas the effect on MMSE was not statistically significant for the majority of estimators(i.e., confidence intervals included zero; Figure 2). Additionally, we observed a slightly negative but non-significant effect on CDR, while ATE estimates for GDS were uniformly non-significant. Moreover, we identified a clearly negative effect on motor function as measured by the FAQ score, with ATEs greater than −1 for 4 out of 5 estimators, corresponding to an average increase of approximately 3 points over three years on the FAQ scale in the treated group compared with controls. Although this effect does not reach statistical significance for all estimators, it is consistently close to the significance threshold.

Overall, these results suggest that ChEIs may be associated with a deleterious effect on cognitive and functional outcomes, although not consistently supported across all estimators.

##### Short-term Outcome

In contrast, short-term results showed substantial heterogeneity in both magnitude and statistical significance across outcomes and estimators (Figure 3). We observed slightly deleterious point estimates for MOCA and MMSE. However, only 2 out of 5 estimators yielded statistically significant effects, while the remaining estimators produced non significative and sometimes even positive point estimates. Additionally, small negative effects were observed for the motor score FAQ, as well as for GDS and CDR, but these effects were not statistically significant for FAQ and reached significance for only 2 out of 5 estimators for GDS and CDR. This limited consistency across estimators complicates the interpretation of short-term treatment effects.

Overall, the lack of robustness across estimators restricts firm conclusions regarding short-term outcomes, despite suggestive trends toward negative effects on motor function (FAQ) and slight negative effects on cognitive outcomes (MOCA and MMSE), which are directionally consistent with the long-term findings.

The placebo refutation analysis yielded an estimated ATE centered around zero (see Figure 11 in Appendix), indicating no spurious treatment effect.

#### Average Treatment Effect in Women and Men

Figure 4 compares long-term ATEs across all outcomes between women and men using the causal forest estimator. For cognitive outcomes, ChEIs appear to have slightly more deleterious effects in men than in women, with ATE point estimates of −0.52 for women versus −0.83 for men for MOCA, and −0.04 for women versus −0.09 for men for MMSE, and ATE point estimates of −0.38 versus −0.59 for men. For motor function as measured by FAQ, treatment effects are similar across sexes, while effects on GDS are not statistically significant in either group.

**Figure 4:**
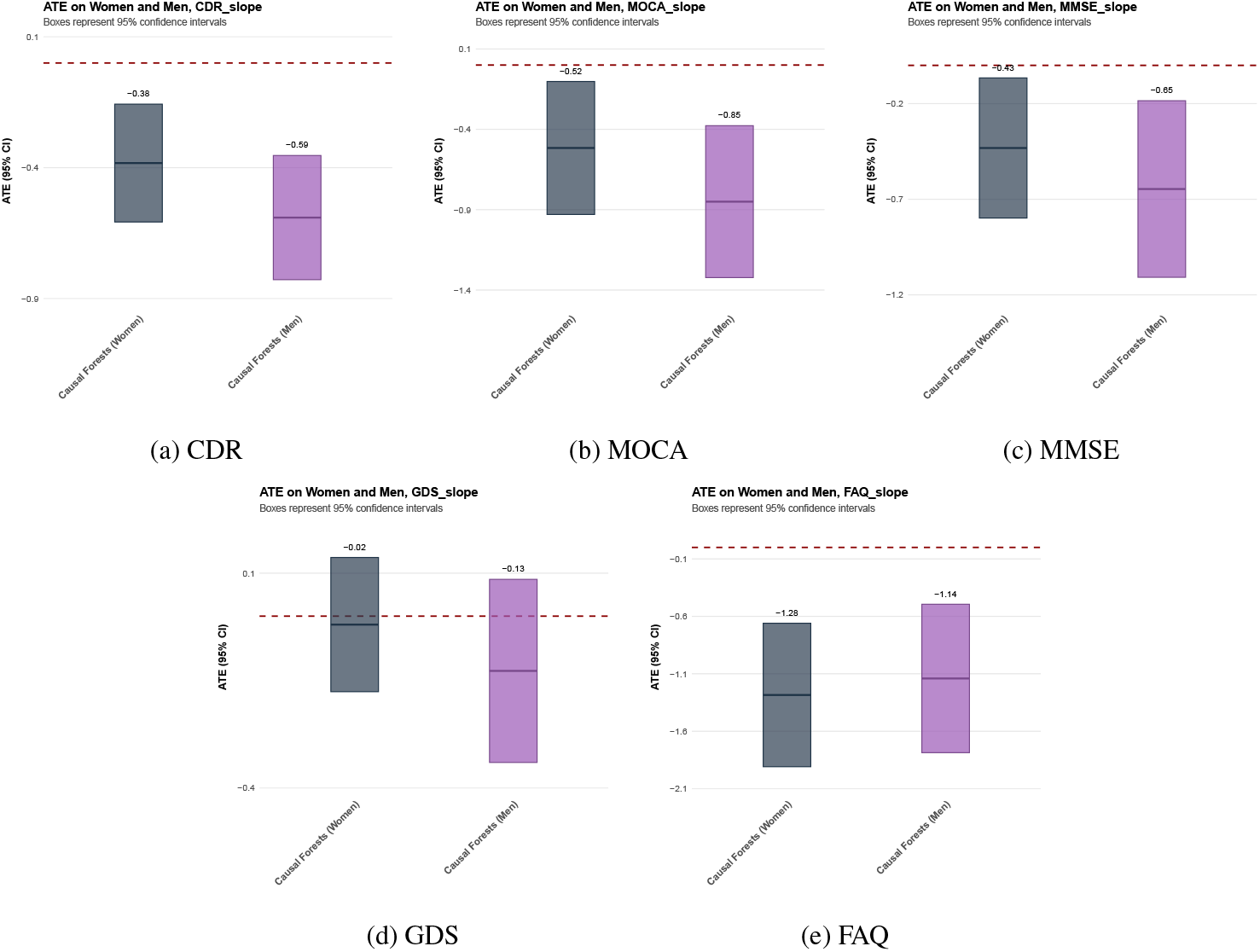
Average Treatment Effects in women and men on multiple outcomes on the long-term with causal forests

Overall, these sex-stratified results suggest that sex may act as a small moderator of treatment effects, with differential patterns observed across cognitive and motor-related outcomes.

#### Subgroup identification

Subgroup identification was applied to the population on the long-term outcome, CDR. To reduce overfitting of the policy tree, we chose a maximum depth of 3. The resulting tree indicated that the most decisive variables for recommending treatment for long-term were BMI and clinical scores at baseline MMSE and FAQ.

As shown in Figure 5, no statistically significant differences emerged: the estimated policy value closely mirrored the ATE. This result admits a two-fold interpretation. First, it suggests limited treatment effect heterogeneity in the data, such that individualized treatment assignment offers little improvement over a uniform policy. Second, even if some heterogeneity is present, the sample size may be insufficient to reliably learn an optimal policy capable of exploiting it, especially if treatment heterogeneity is small.

**Figure 5:**
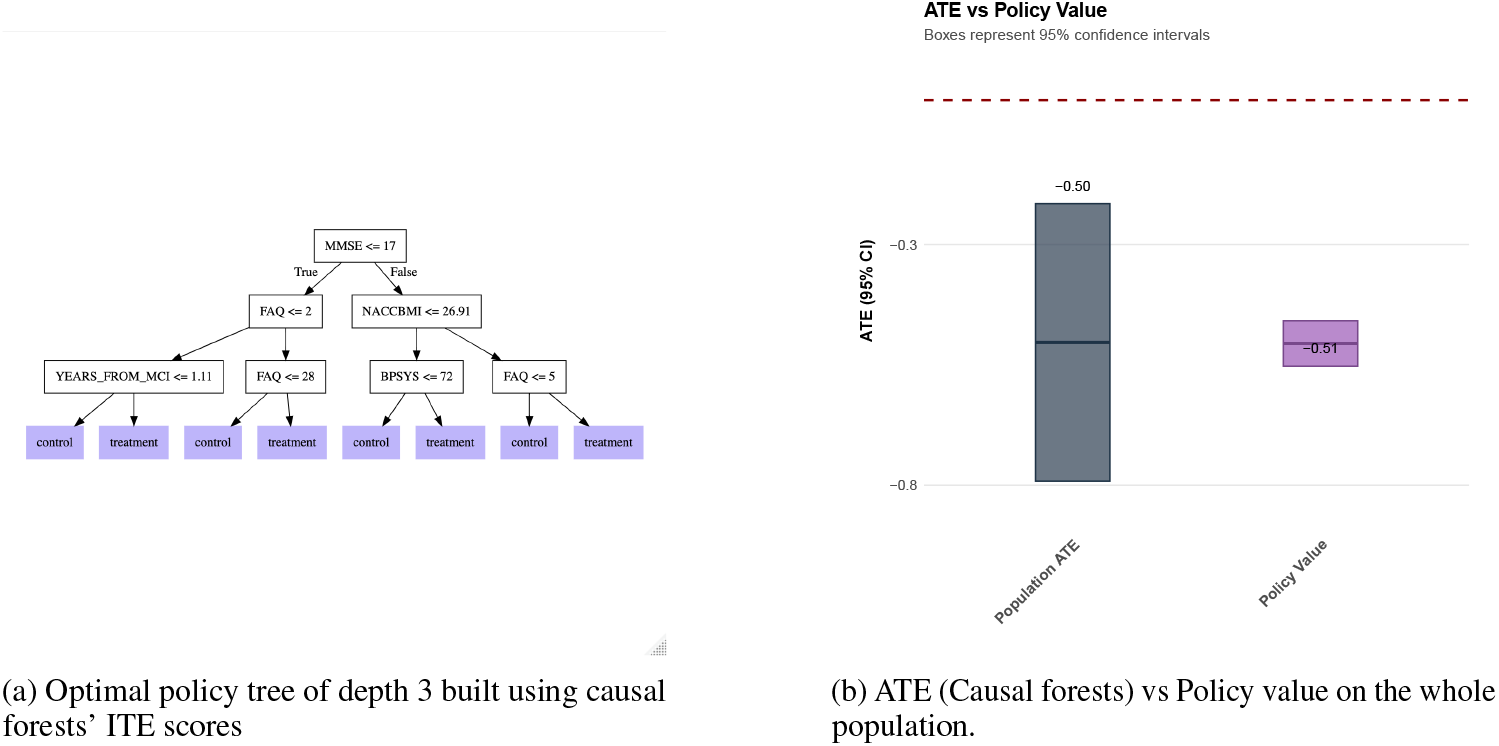
Heterogeneity analysis with policy tree

**Figure 6:**
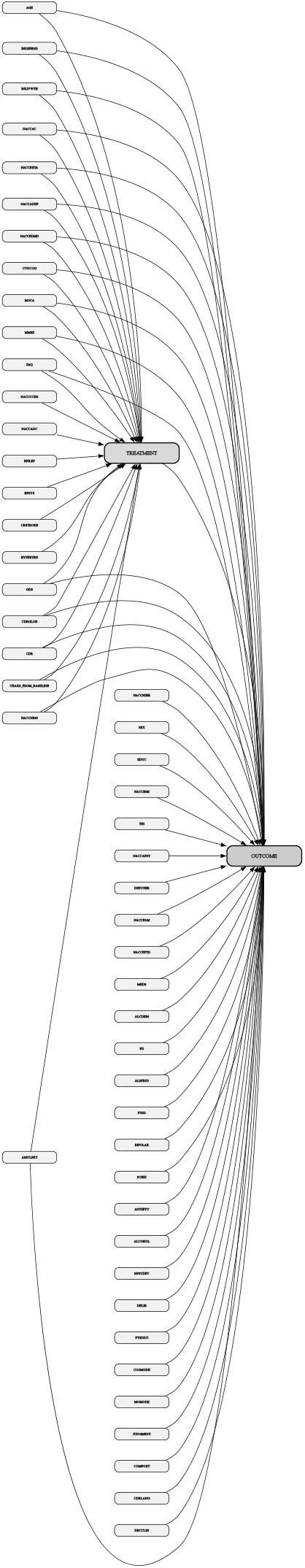
Causal diagram used in the analysis. All variables shown are derived from the National Alzheimer’s Coordinating Center (NACC) dataset. Definitions and coding of variables follow the NACC data dictionary (https://naccdata.org/data-dictionary).

**Figure 7:**
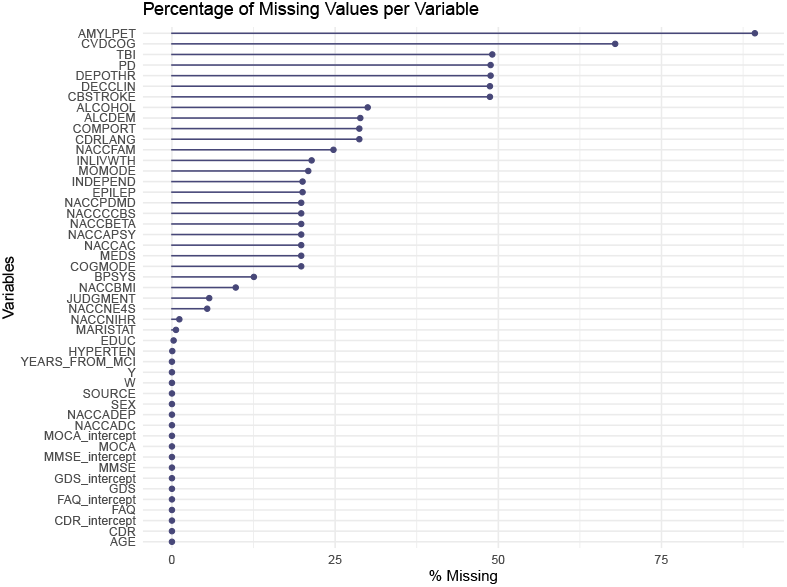
Missing value percentages

**Figure 8:**
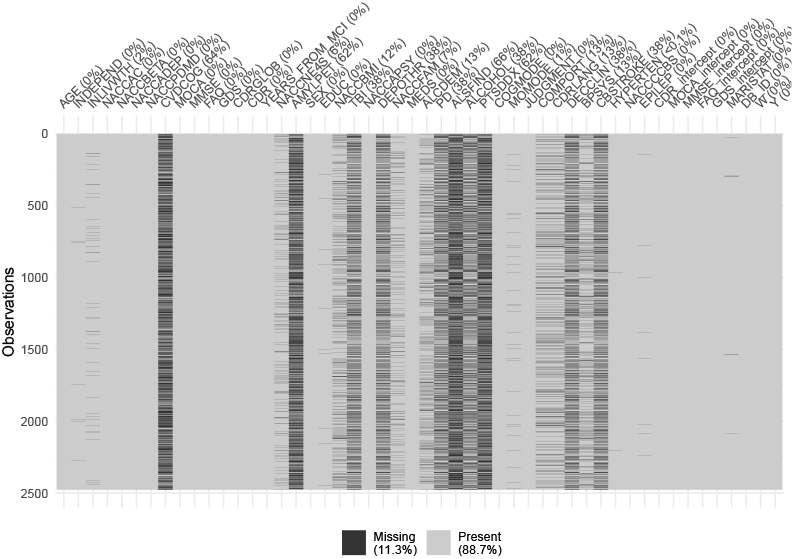
Missing value table

**Figure 9:**
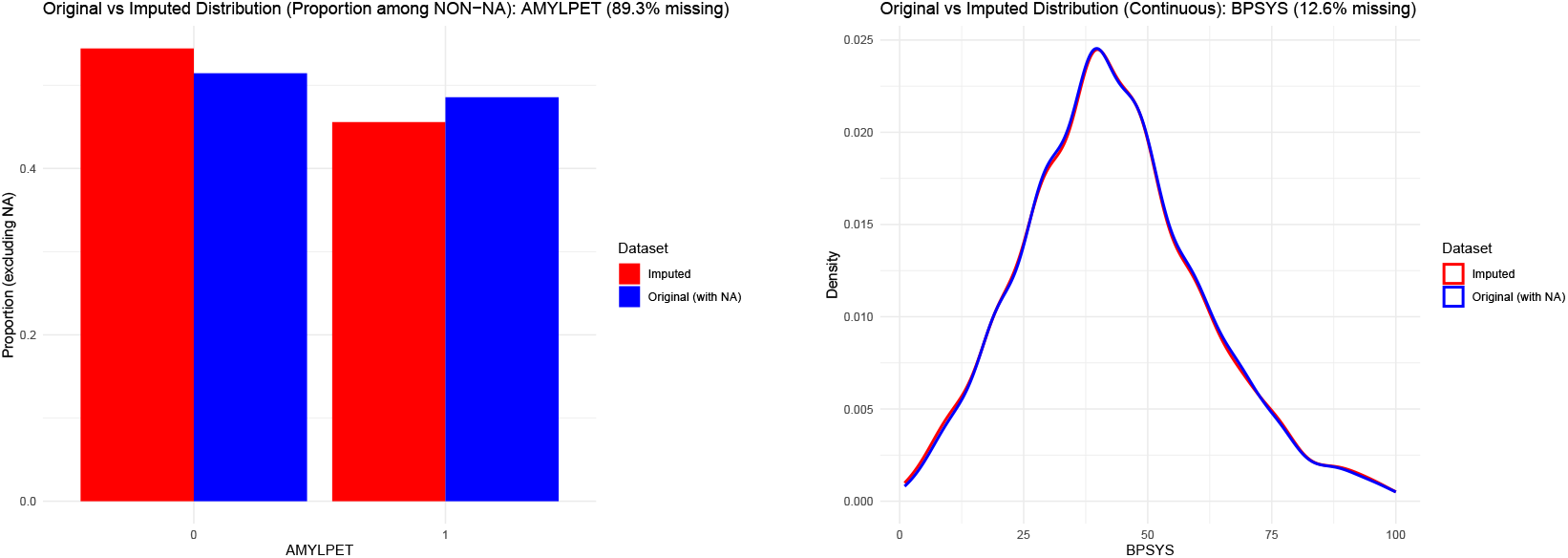
Value distributions of different variables on imputed and non-imputed datasets. Examples of two variables: a categorical one (left) for which we obtain chart plots, and continuous ones (right) with estimated densities.

**Figure 10:**
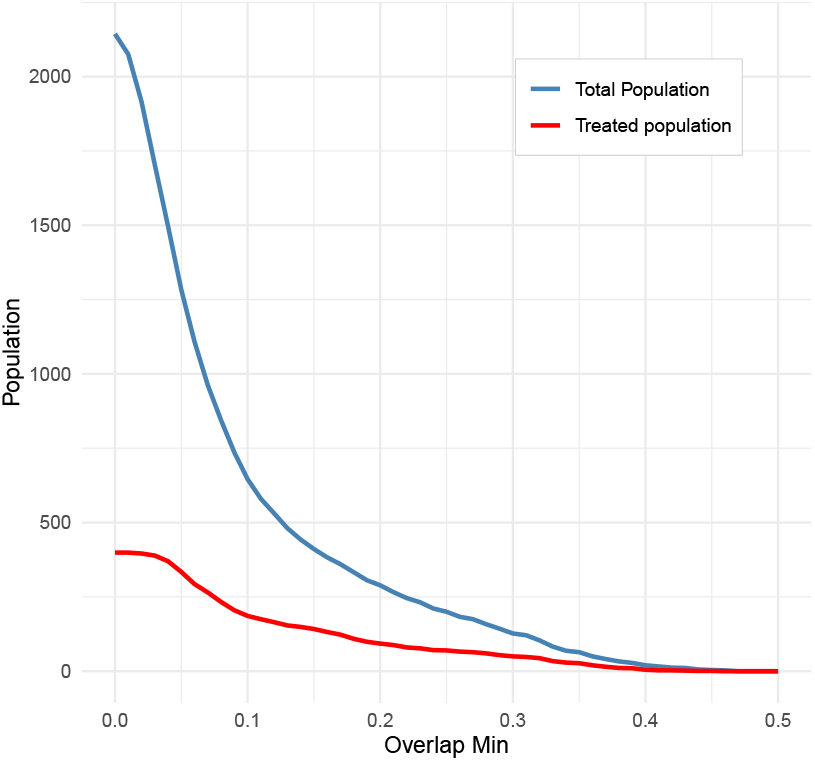
Populations corresponding to overlap truncations

**Figure 11:**
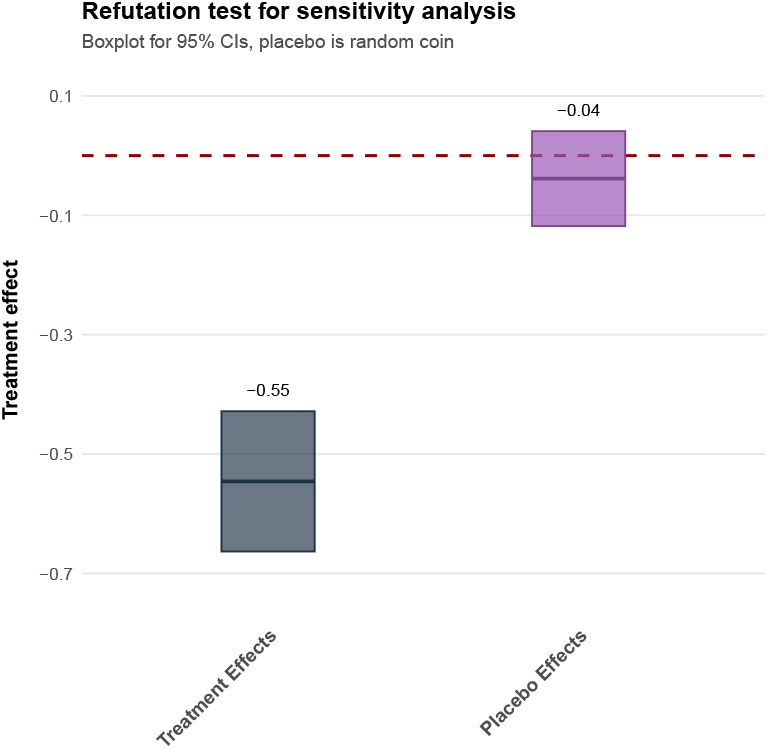
Average Treatment Effects for the CDR outcome, for the treatment ChEI and for a virtual randomly assigned treatment

### 4 Discussion

#### 4.1 Key findings and clinical interpretation

Overall, our results indicated that the effects of ChEIs differ across outcome domains and time horizons. In the short term, treatment effects were heterogeneous and generally small, with no consistent evidence of cognitive or functional benefit. In the long term, results were more consistent across estimators, pointing to a small negative or null effect on cognitive performance most notably for MOCA and a stronger negative effect on functional outcomes. These findings suggested that ChEIs were unlikely to provide sustained cognitive or functional benefits on the long term. These findings are consistent with recent observational studies based on the ADNI and NACC cohorts, which reported an association between ChEI use and accelerated cognitive decline [21, 23]. Notably, these studies did not examine functional outcomes such as the FAQ, making the negative motor effects identified in our analysis a novel contribution.

The clinical relevance of our results nevertheless warrants careful consideration, as part of the existing literature reports beneficial effects of ChEIs on cognitive outcomes and less frequently on functional outcomes, which have received comparatively less attention over both short and long-term follow-up [10, 18, 11, 17, 40]. Several factors may account for discrepancies with prior studies. First, in cohorts such as ADNI and NACC, amyloid status is frequently unavailable, raising the possibility of diagnostic heterogeneity, which may obscure true treatment effects. This concern is underscored by evidence of misdiagnosis rates of up to 35% in early AD when biomarkers are not used [41], supporting the use of biomarker-confirmed amyloid positivity as a standard inclusion criterion, even for non-immunotherapy interventions. Second, our study focused on very early-stage patients (CDR 0 or 0.5), a group known to exhibit substantial heterogeneity in disease trajectories, with some individuals declining rapidly while others remain stable [42, 43]. Such variability may affect estimated treatment effects in both treated and untreated groups. Together, these considerations highlight the need for replication in independent, well-characterized cohorts.

The biological plausibility of these null or adverse findings on the cognitive function merits consideration. ChEIs enhance cholinergic neurotransmission by inhibiting acetylcholinesterase, but MCI patients may have relatively intact cholinergic systems compared to those with established AD dementia. Cholinergic augmentation in the absence of substantial cholinergic deficit may provide limited benefit or even prove detrimental through receptor desensitization or disruption of finely tuned neurotransmitter balance.

Another objective of this study was to assess sex-specific differences in treatment response and to identify potential responder subgroups. Sex-stratified analyses suggested modest differences in treatment effects, with slightly more negative cognitive effects observed in men than in women, and conversely more favorable functional effects in men. These findings indicated that sex may act as a weak modifier of treatment response. The existing literature on sex-specific responses to ChEIs is mixed, with prior studies reporting greater benefit in men, greater benefit in women, or no significant sex-related differences across different ChEI agents and study populations [6]. While documented sex-related differences in cholinergic neurotransmission [4] provide biological plausibility for differential responses, the magnitude of sex effects observed in our study was small, limiting their immediate clinical relevance.

In addition, our analyses did not identify any statistically significant responder subgroups, and policy learning algorithms failed to uncover clinically meaningful treatment rules. Given existing evidence of heterogeneity in treatment response, this absence of detectable subgroups may reflect limited statistical power, as causal machine learning methods for heterogeneity estimation typically require large sample sizes.

#### 4.2 Limitations and future directions

Several limitations arouse from the inherent challenges of applying causal inference to observational data. First, the validity of the estimated treatment effects depended on causal assumptions that may not be fully satisfied, and residual bias cannot be entirely excluded. In particular, preprocessing steps required to support valid causal assumptions implying baseline alignment, overlap trimming, and censoring at treatment switches, which substantially reduced the analytic sample, thereby limiting statistical power, especially for the detection of small or heterogeneous effects. Although we adjusted for a wide range of demographic, clinical, and genetic factors, residual confounding from unmeasured variables remains possible and may still bias estimated treatment effects. Second, outcomes were summarized as longitudinal slopes, which may oversimplify disease trajectories that could be better captured using more flexible statistical models. In addition, our analyses were restricted to cognitive and functional scales that are subject to ceiling effects, potentially obscuring benefits in quality of life, caregiver burden, or specific cognitive domains. The inclusion of patient-reported outcomes and biomarker-based endpoints could provide a more comprehensive assessment of treatment effects. Finally, ChEI exposure was modeled as a binary variable, without accounting for dose; approaches incorporating time-varying exposure such as marginal structural models would allow a more accurate representation of treatment dynamics including treatment switches and dose adjustments.

The primary value of this work lies in establishing a generalizable framework for estimating treatment effects and HTE in observational data. Having validated our methods with ChEIs, we can now apply them to treatments where evidence is more limited or heterogeneity more plausible. Memantine represents a compelling next application. As an N-methyl-D-aspartate (NMDA) receptor antagonist with distinct mechanisms from ChEIs, it may exhibit different heterogeneity patterns. Sex differences in glutamatergic neurotransmission suggest plausible mechanisms for differential responses [44]. Anti-amyloid monoclonal antibodies (lecanemab, donanemab) present urgent applications. These disease-modifying therapies show modest average benefits but carry substantial risks and costs, making responder identification critical. RCT subgroup analyses hint at differential efficacy by APOE status, amyloid burden, and tau pathology [45]. As RWD accumulates, our framework could rigorously characterize which patients benefit most. Moreover, this framework enables comparative effectiveness research and optimal treatment assignement. One could estimate individual level outcomes under different strategies, e.g., monotherapy, combination, sequential therapy, and propose personalized treatment rules via policy learning.

#### 4.3 Conclusion

This study developed and validated a comprehensive causal-ML framework to estimate treatment effects and explore HTE in AD using observational data while accounting for confounding, missing data, and selection bias. We explicitly addressed treatment selection bias through the integration of clinical expert knowledge and rigorous causal inference methods designed to move beyond correlation toward causal interpretation in observational cohorts. Key components included expert-guided causal graph design, the use of multiple complementary estimators validated through propensity score diagnostics, and sensitivity analyses. Importantly, our approach evaluated the effects of ChEIs on multiple outcomes, both cognitive and functional outcomes, and across different time horizons, both short and long-term.

Moreover, applying this framework, we showed that ChEIs do not confer sustained long-term benefits on cognitive and functional outcomes. In addition, the absence of clearly identifiable responder subgroups highlighted the challenges of detecting HTEs in observational settings and suggested that substantially larger sample sizes may be required to achieve sufficient statistical power and clinical relevance.

Finally, this framework provides a methodological foundation for future causal analyses in neurodegenerative disease research. As disease-modifying therapies are increasingly introduced and biomarker data become more widely available, robust methods for predicting individual treatment responses will be essential. Future work should extend this approach to additional therapies and incorporate biomarker-based endpoints to further enhance precision treatment strategies in AD.

## Data Availability

All data produced in the present work are contained in the manuscript.

## Acknowledgments

We thank Iona Findji for her valuable assistance in the design of questionnaires for clinicians, and Julie Josse for her methodological guidance throughout this study.

## Consent statement

ADNI was approved by the Institutional Review Boards of all participating institutions. All ADNI participants provided written informed consent according to the Declaration of Helsinki before study enrollment.

The NACC database is funded by NIA/NIH Grant U24 AG072122. NACC data are contributed by the NIA-funded ADRCs: P30 AG062429 (PI James Brewer, MD, PhD), P30 AG066468 (PI Oscar Lopez, MD), P30 AG062421 (PI Bradley Hyman, MD, PhD), P30 AG066509 (PI Thomas Grabowski, MD), P30 AG066514 (PI Mary Sano, PhD), P30 AG066530 (PI Helena Chui, MD), P30 AG066507 (PI Marilyn Albert, PhD), P30 AG066444 (PI David Holtzman, MD), P30 AG066518 (PI Lisa Silbert, MD, MCR), P30 AG066512 (PI Thomas Wisniewski, MD), P30 AG066462 (PI Scott Small, MD), P30 AG072979 (PI David Wolk, MD), P30 AG072972 (PI Charles DeCarli, MD), P30 AG072976 (PI Andrew Saykin, PsyD), P30 AG072975 (PI Julie A. Schneider, MD, MS), P30 AG072978 (PI Ann McKee, MD), P30 AG072977 (PI Robert Vassar, PhD), P30 AG066519 (PI Frank LaFerla, PhD), P30 AG062677 (PI Ronald Petersen, MD, PhD), P30 AG079280 (PI Jessica Langbaum, PhD), P30 AG062422 (PI Gil Rabinovici, MD), P30 AG066511 (PI Allan Levey, MD, PhD), P30 AG072946 (PI Linda Van Eldik, PhD), P30 AG062715 (PI Sanjay Asthana, MD, FRCP), P30 AG072973 (PI Russell Swerdlow, MD), P30 AG066506 (PI Glenn Smith, PhD, ABPP), P30 AG066508 (PI Stephen Strittmatter, MD, PhD), P30 AG066515 (PI Victor Henderson, MD, MS), P30 AG072947 (PI Suzanne Craft, PhD), P30 AG072931 (PI Henry Paulson, MD, PhD), P30 AG066546 (PI Sudha Seshadri, MD), P30 AG086401 (PI Erik Roberson, MD, PhD), P30 AG086404 (PI Gary Rosenberg, MD), P20 AG068082 (PI Angela Jefferson, PhD), P30 AG072958 (PI Heather Whitson, MD), P30 AG072959 (PI James Leverenz, MD).

## Appendix

### 4.4 Methods

#### Average Treatment Effect & Causal Assumptions

One can observe the outcome given whether the patient takes the treatment or not, and the other unobserved outcome is called the potential outcome (or counterfactual outcome). Then we can estimate the ATE using its definition:

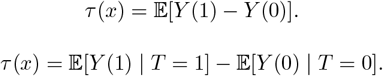

To estimate the CATE, we have to establish the following assumptions:

##### Ignorability

Treatment assignment is independent of the outcome conditioned on covariates, which is

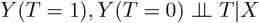

##### Positivity

For every possible value of the covariates *X*, each treatment level has a strictly positive probability of being assigned. Formally, it can be written as:

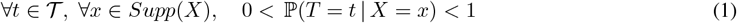

##### SUTVA

Potential outcomes do not vary given treatments assigned to others (no interference), and treatment level or dosage is unique (consistency).

The assumptions introduced above allow us to identify the CATE. By definition:

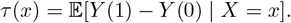

#### Covariate balancing & Trimming

Violations of the positivity assumption arise when certain covariate profiles almost perfectly predict treatment assignment, leading to extreme inverse-probability weights and unstable estimators.

To mitigate this issue, we implemented a trimming procedure based on the estimated propensity score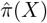. Specifically, individuals with

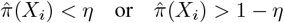

were excluded from the analytic sample, for some threshold *η >* 0. The value of *η* was selected empirically to balance sufficient overlap with adequate retention of treated individuals. All subsequent causal analyses were performed on this restricted population.

Evaluating covariate balance between treated and untreated groups is essential to validate propensity score based causal analyses. Following standard practice, we quantified balance using the *Standardized Mean Difference* (SMD), defined for a given covariate *X*_*j*_ as

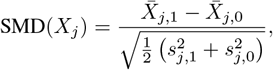

where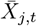 and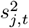 denote the sample mean and variance in treatment group *t* ∈ {0, 1}.

An absolute SMD below a prespecified threshold (typically 0.1) is commonly interpreted as evidence of adequate balance. To assess the quality of the propensity score model, covariate balance was evaluated both *before* and *after* applying the propensity scores. This ensured that the weighting procedure effectively reduced systematic differences between groups. All causal estimations in this study were performed only after confirming that post-weighting SMDs met an acceptable covariate balancing.

### 4.5 Results

#### 4.5.1 Study Population

#### 4.5.2 Variables

#### 4.5.3 Imputation of Missing Values

#### 4.5.4 Covariate Balancing

#### 4.5.5 Placebo Refutation Analysis

The placebo refutation analysis yielded an estimated ATE centered around zero (see Figure 11), indicating no spurious treatment effect.

## References

“2024 Alzheimer’s disease facts and figures” (2024). In: Alzheimer’s & Dementia 20.5, pp. 3708–3821.

Duara, R. and W. Barker (2022). “Heterogeneity in Alzheimer’s Disease Diagnosis and Progression Rates: Implications for Therapeutic Trials”. In: Neurotherapeutics 19.1, pp. 8–25.

Ferretti, M. T., M. F. Iulita, E. Cavedo, P. A. Chiesa, A. Schumacher Dimech, A. Santuccione Chadha, F. Baracchi, H. Girouard, S. Misoch, E. Giacobini, H. Depypere, and H. Hampel (2018). “Sex differences in Alzheimer disease — the gateway to precision medicine”. In: Nature Reviews Neurology 14.8, pp. 457–469.

Giacobini, E. and G. Pepeu (2018). “Sex and Gender Differences in the Brain Cholinergic System and in the Response to Therapy of Alzheimer Disease with Cholinesterase Inhibitors”. In: Current Alzheimer Research 15.11, pp. 1077– 1084.

Climacosa, F. M. M., E. D. B. Ornos, N. C. L. L. Gapaz, M. G. R. Guantia, J. M. C. Cruz, R. V. M. Manalo, M. L. Yu, A. P. Qureshi, A. C. Carampel, J. L. B. Asis, J. C. B. Reyes, A. B. Dacasin, and V. M. M. Anlacan (2025). “The role of genetic polymorphisms on drug response in Alzheimer’s disease: a systematic review”. In: BMC Medical Genomics 18, p. 154.

Lynch, M. A. (2024). “A case for seeking sex-specific treatments in Alzheimer’s disease”. In: Frontiers in Aging Neuroscience 16, p. 1346621.

Buckley, R. F., J. Gong, and M. Woodward (2023). “A Call to Action to Address Sex Differences in Alzheimer Disease Clinical Trials”. In: JAMA Neurology 80.8, pp. 769–770.

Abécassis, J., É. Dumas, J. Alberge, and G. Varoquaux (2025). “From Prediction to Prescription: Machine Learning and Causal Inference for the Heterogeneous Treatment Effect”. In: Annual Review of Biomedical Data Science 8.1, pp. 381–404.

Sanchez, P., J. P. Voisey, T. Xia, H. I. Watson, A. Q. O’Neil, and S. A. Tsaftaris (2022). “Causal machine learning for healthcare and precision medicine”. In: Royal Society Open Science 9.8, p. 220638.

Rogers, S. L., M. R. Farlow, R. S. Doody, R. Mohs, L. T. Friedhoff, and Donepezil Study Group* (1998). “A 24-Week, Double-Blind, Placebo-Controlled Trial of Donepezil in Patients with Alzheimer’s Disease”. In: Neurology 50.1, pp. 136–145.

Feldman, H., S. Gauthier, J. Hecker, B. Vellas, P. Subbiah, E. Whalen, and the Donepezil MSAD Study Investigators Group* (2001). “A 24-week, randomized, double-blind study of donepezil in moderate to severe Alzheimer’s disease”. In: Neurology 57.4, pp. 613–620.

Lanctôt, K. L., N. Herrmann, K. K. Yau, L. R. Khan, B. A. Liu, M. M. LouLou, and T. R. Einarson (2003). “Efficacy and safety of cholinesterase inhibitors in Alzheimer’s disease: a meta-analysis”. en. In: CMAJ 169.6, pp. 557–564.

Birks, J. (2006). “Cholinesterase Inhibitors for Alzheimer’s Disease”. In: The Cochrane Database of Systematic Reviews 2006.1, p. CD005593.

Hansen, R. A., G. Gartlehner, A. P. Webb, L. C. Morgan, C. G. Moore, and D. E. Jonas (2008). “Efficacy and safety of donepezil, galantamine, and rivastigmine for the treatment of Alzheimer’s disease: A systematic review and meta-analysis”. In: Clinical Interventions in Aging 3.2, pp. 211–225.

Santoro, A., P. Siviero, N. Minicuci, E. Bellavista, M. Mishto, F. Olivieri, F. Marchegiani, A. M. Chiamenti, L. Benussi, R. Ghidoni, B. Nacmias, S. Bagnoli, A. Ginestroni, O. Scarpino, E. Feraco, W. Gianni, G. Cruciani, R. Paganelli, A. Di Iorio, M. Scognamiglio, L. M. E. Grimaldi, C. Gabelli, S. Sorbi, G. Binetti, G. Crepaldi, and C. Franceschi (2010). “Effects of Donepezil,Galantamine and Rivastigmine in 938 Italian Patients with Alzheimer’s Disease”. In: CNS Drugs 24.2, pp. 163–176.

Winblad, B., K. Engedal, H. Soininen, F. Verhey, G. Waldemar, A. Wimo, A.-L. Wetterholm, R. Zhang, A. Haglund, P. Subbiah, and the Donepezil Nordic Study Group (2001). “A 1-year, randomized, placebo-controlled study of donepezil in patients with mild to moderate AD”. In: Neurology 57.3, pp. 489–495.

Rösler, M., W. Retz, P. Retz-Junginger, and H. J. Dennler (1999). “Effects of Two-Year Treatment with the Cholinesterase Inhibitor Rivastigmine on Behavioural Symptoms in Alzheimer’s Disease”. en. In: Behavioural Neurology 11.4, p. 168023.

Rountree, S. D., W. Chan, V. N. Pavlik, E. J. Darby, S. Siddiqui, and R. S. Doody (2009). “Persistent treatment with cholinesterase inhibitors and/or memantine slows clinical progression of Alzheimer disease”. en. In: Alzheimers Res Ther 1.2, p. 7.

Xu, H., S. Garcia-Ptacek, L. Jönsson, A. Wimo, P. Nordström, and M. Eriksdotter (2021). “Long-Term Effects of Cholinesterase Inhibitors on Cognitive Decline and Mortality”. In: Neurology 96.17, e2220–e2230.

Schneider, L. S., P. S. Insel, M. W. Weiner, and Alzheimer’s Disease Neuroimaging Initiative (2011). “Treatment With Cholinesterase Inhibitors and Memantine of Patients in the Alzheimer’s Disease Neuroimaging Initiative”. In: Archives of Neurology 68.1, pp. 58–66.

Han, J.-y., L. M. Besser, C. Xiong, W. A. Kukull, and J. C. Morris (2019). “Cholinesterase Inhibitors May Not Benefit Mild Cognitive Impairment and Mild Alzheimer Disease Dementia”. en-US. In: Alzheimer Disease & Associated Disorders 33.2, p. 87.

Pyun, J.-M., Y. H. Park, M. J. Kang, and S. Kim (2024). “Cholinesterase inhibitor use in amyloid PET-negative mild cognitive impairment and cognitive changes”. en. In: Alzheimer’s Research & Therapy 16.1, p. 210.

Liu, W., Y. Li, W. Qin, X. Wang, W. Li, Y. Li, A. D. N. Initiative, and J. Jia (2025). “The impact of cholinesterase inhibitors on cognitive trajectories in mild cognitive impairment patients based on amyloid beta status”. en. In: Alzheimer’s & Dementia 21.5, e70193.

Blanco-Silvente, L., X. Castells, M. Saez, M. A. Barceló, J. Garre-Olmo, J. Vilalta-Franch, and D. Capellà (2017). “Discontinuation, Efficacy, and Safety of Cholinesterase Inhibitors for Alzheimer’s Disease: a Meta-Analysis and Meta-Regression of 43 Randomized Clinical Trials Enrolling 16 106 Patients”. In: The International Journal of Neuropsychopharmacology 20.7, pp. 519–528.

Li, D.-D., Y.-H. Zhang, W. Zhang, and P. Zhao (2019). “Meta-Analysis of Randomized Controlled Trials on the Efficacy and Safety of Donepezil, Galantamine, Rivastigmine, and Memantine for the Treatment of Alzheimer’s Disease”. In: Frontiers in Neuroscience 13, p. 472.

Morris, J. C., S. Weintraub, H. C. Chui, J. Cummings, C. DeCarli, S. Ferris, N. L. Foster, D. Galasko, N. Graff-Radford, E. R. Peskind, D. Beekly, E. M. Ramos, and W. A. Kukull (–2006). “The Uniform Data Set (UDS): Clinical and Cognitive Variables and Descriptive Data From Alzheimer Disease Centers”. In: Alzheimer Disease & Associated Disorders 20.4, p. 210.

Beekly, D. L., E. M. Ramos, W. W. Lee, W. D. Deitrich, M. E. Jacka, J. Wu, J. L. Hubbard, T. D. Koepsell, J. C. Morris, W. A. Kukull, and T. N. A. D. Centers (–2007). “The National Alzheimer’s Coordinating Center (NACC) Database: The Uniform Data Set”. In: Alzheimer Disease & Associated Disorders 21.3, p. 249.

Weintraub, S., D. Salmon, N. Mercaldo, S. Ferris, N. R. Graff-Radford, H. Chui, J. Cummings, C. DeCarli, N. L. Foster, D. Galasko, E. Peskind, W. Dietrich, D. L. Beekly, W. A. Kukull, and J. C. Morris (–2009). “The Alzheimer’s Disease Centers’ Uniform Data Set (UDS): The Neuropsychologic Test Battery”. In: Alzheimer Disease & Associated Disorders 23.2, p. 91.

Mueller, S. G., M. W. Weiner, L. J. Thal, R. C. Petersen, C. Jack, W. Jagust, J. Q. Trojanowski, A. W. Toga, and L. Beckett (2005). “The Alzheimer’s Disease Neuroimaging Initiative”. In: Neuroimaging Clinics 15.4, pp. 869–877.

Weiner, M. W., P. S. Aisen, C. R. Jack, W. J. Jagust, J. Q. Trojanowski, L. Shaw, A. J. Saykin, J. C. Morris, N. Cairns, L. A. Beckett, A. Toga, R. Green, S. Walter, H. Soares, P. Snyder, E. Siemers, W. Potter, P. E. Cole, and M. Schmidt (2010). “The Alzheimer’s Disease Neuroimaging Initiative: Progress Report and Future Plans”. In: Alzheimer’s & Dementia 6.3, 202–211.e7.

Morris, J. C. (1993). “The Clinical Dementia Rating (CDR)”. In: Neurology 43.11, 2412-2412–a.

Rubin, D. B. (1974). “Estimating causal effects of treatments in randomized and nonrandomized studies.” In: Journal of educational Psychology 66.5, p. 688.

Rosenbaum, P. R. (2002). “Observational studies”. In: Observational studies. Springer, pp. 1–17.

Wager, S. (2024). Causal inference: A statistical learning approach.

Chernozhukov, V., D. Chetverikov, M. Demirer, E. Duflo, C. Hansen, W. Newey, and J. Robins (2018). Double/debiased machine learning for treatment and structural parameters.

Athey, S., J. Tibshirani, and S. Wager (2019). “Generalized Random Forests”. In: The Annals of Statistics 47.2, pp. 1148– 1178.

Nie, X. and S. Wager (2021). “Quasi-oracle estimation of heterogeneous treatment effects”. In: Biometrika 108.2, pp. 299–319.

Efron, B. (1981). “Nonparametric estimates of standard error: the jackknife, the bootstrap and other methods”. In: Biometrika 68.3, pp. 589–599.

Sverdrup, E., A. Kanodia, Z. Zhou, S. Athey, and S. Wager (2020). “policytree: Policy learning via doubly robust empirical welfare maximization over trees”. In: Journal of Open Source Software 5.50, p. 2232.

Xu, Y., J. S. Kim, L. K. Hummers, A. A. Shah, and S. L. Zeger (2024). “Causal Inference Using Multivariate Generalized Linear Mixed-Effects Models”. In: Biometrics 80.3, ujae100.

Rabinovici, G. D., C. Gatsonis, C. Apgar, K. Chaudhary, I. Gareen, L. Hanna, J. Hendrix, B. E. Hillner, C. Olson, O. H. Lesman-Segev, J. Romanoff, B. A. Siegel, R. A. Whitmer, and M. C. Carrillo (2019). “Association of Amyloid Positron Emission Tomography With Subsequent Change in Clinical Management Among Medicare Beneficiaries With Mild Cognitive Impairment or Dementia”. In: JAMA 321.13, pp. 1286–1294.

Jutten, R. J., S. A. Sikkes, W. M. Van der Flier, P. Scheltens, P. J. Visser, B. M. Tijms, and for the Alzheimer’s Disease Neuroimaging Initiative (2021). “Finding Treatment Effects in Alzheimer Trials in the Face of Disease Progression Heterogeneity”. In: Neurology 96.22, e2673–e2684.

Wang, G., Y. Li, E. McDade, C. Xiong, S. M. Hartz, R. J. Bateman, J. C. Morris, and L. S. Schneider (2025). “Clinical progression on CDR-SB©: Progression-free time at each 0.5 unit level in dominantly inherited and sporadic Alzheimer’s disease populations”. In: Alzheimer’s & Dementia 21.9, e70643.

Wickens, M. M., D. A. Bangasser, and L. A. Briand (2018). “Sex Differences in Psychiatric Disease: A Focus on the Glutamate System”. In: Frontiers in Molecular Neuroscience 11.

Pang, M., A. Gabelle, P. Saha-Chaudhuri, W. Huijbers, A. Gafson, P. M. Matthews, L. Tian, I. Rubino, R. Hughes, C. De Moor, S. Belachew, and C. Shen (2024). “Precision medicine analysis of heterogeneity in individual-level treatment response to amyloid beta removal in early Alzheimer’s disease”. In: Alzheimer’s & Dementia 20.2, pp. 1102–1111.

